# Decrease of heart rate variability during exercise: an index of cardiorespiratory fitness

**DOI:** 10.1101/2021.09.23.21263943

**Authors:** Denis Mongin, Clovis Chabert, Manuel Gomez Extremera, Olivier Hue, Delphine Sophie Courvoisier, Pedro Carpena, Pedro Angel Bernaola Galvan

## Abstract

The present study proposes to measure and quantify the heart rate variability (HRV) changes during effort and to test the capacity of the produced indices to predict cardiorespiratory fitness measures. Therefore, the beat-to-beat cardiac time interval series of 18 adolescent athletes (15.2 ± 2.0 years) measured during maximal graded effort test were detrended using a dynamical first-order differential equation model. Heart rate variability was then calculated as the standard deviation of the detrended RR intervals within successive windows of one minute. The variation of this measure of HRV during exercise is properly adjusted by an exponential decrease of the heart rate. The amplitude and the decay rate of this exponential trend are strongly associated with maximum oxygen consumption, maximal aerobic power, and ventilatory thresholds. It indicates that among athletes with better fitness, HRV has higher values at low heart rate and decreases faster when the heart rate increases during exercise. This analysis, based only on cardiac measurements, provides a promising tool for the study of cardiac measurements generated by portable devices.

## Introduction

The human heart is involved in the response to the energy demand of the body. Its regulation is mainly driven by the subtle balance between the sympathetic and parasympathetic branches of the autonomic nervous system (ANS) (1,2). The activity and relative level of these two cardiac neural systems cause the main dynamical changes of heart rate (HR) in response to external stimulus, and on a shorter time scale, the fluctuations of the heart R-wave to R-wave (RR) time interval known as heart rate variability (HRV). HRV during rest has been shown to be influenced by psychological (3) as well as physiological factors such as age, body mass index, diseases (4), heart functions and heart diseases (5,6), body position (7) and physical fitness (8,9). During physical exercise, HRV dynamics is drastically modified due to the break of the balance between both branches of ANS. The progressive withdrawal of the parasympathetic activity and the subsequent increase of the sympathetic activity causes extensive changes in RR intervals, such as the decrease of their variability measured both in time and frequency domain (8,10–15), the modification of the scaling properties of their linear correlations (16), or even the reduction of their sample entropy (17) and of their nonlinear correlations (18,19).

The decrease of HRV during exercise has been described in most cases, mainly qualitatively, as a function of exercise intensity, measured by the oxygen consumption expressed in percentage of the maximum oxygen consumption (%VO_2_max) (7,8,10,12). Lewis and co-authors (11) modeled the decrease of the absolute high frequency (HF) and low frequency (LF) power of the RR series spectrum as an exponential decay of the work load. They suggest that the decay constant of such exponential regression could be an indicator of the physical capacity. To our knowledge, no other study has tried to demonstrate this assertion nor to use this promising index. That could be explained by three main reasons, as follows: i) the inherent need to remove the main RR decrease due to the metabolic response to the energy expenditure during exercise to obtain the detrended RR series, that is the beat-to-beat variability commonly designed as HRV; ii) The technicity and the variety of the HRV characterizations, as illustrated by the variety of spectral measurements used and by the sometimes contradictory results they provide (20,21); iii) the use of the mechanical work rate during exercise as the independent variable of the exponential model, which could restrict the use of such approach only on RR series recorded during effort (HRV before and after effort are associated to the same power value: 0 Watt) and may be protocol dependent.

In the present retrospective analysis, we aimed at testing the ability of an improved and extended approach of the HRV decay time to predict cardiorespiratory fitness (CRF).

## Methods

Each individual RR series was first detrended to obtain a stationary RR series. The detrending has been performed by a tested and characterized dynamical model based on simple physiological considerations [22–24] and free from ad-hoc parameters (see “detrending” subsection). HRV during each individual effort was then quantified by the standard deviation of the detrended RR intervals (SDRR) calculated on adjacent windows of one minute (corresponding to each effort step during effort, see “Heart rate variability” subsection). The mean HR, the mean work intensity %VO_2_max and the mean mechanical power were calculated in the same windows. The analysis was then divided in two parts.

In a first part, three models describing the evolution of the HRV during graded effort test (GET) were compared: the first one represents HRV as an exponential decay of HR, the second one as an exponential decay of the exerted power, and the last one as an exponential decay of the work intensity. More precisely, these models were operationalized as follow:

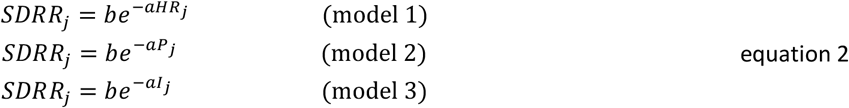

where *HR*_*j*_ is the mean *HR, P*_*j*_ the mean mechanical power exerted and *I*_*j*_ the mean work intensity calculated in same windows *j* as the *SDRR*_*j*_. *a* is the HRV decay rate, and *b* the HRV intercept, i.e. the SDRR corresponding to a hypothetical null heart rate, mechanical power or work intensity respectively. These three models have been implemented by performing least squares nonlinear regressions on the ensemble of the SDRR measurements covering the entire exercise test (measures before the GET and during the recovery are included), and on those during the effort only. They were then compared using Akaike information criterion (AIC) and Bayesian Information criterion (BIC).

In a second part, we used model 1 on each individual SDRR series and tested whether the estimated decrease of HRV as a function of HR is correlated with CRF. Therefore, we performed the least squares nonlinear regression of model 1 (equation 2) for each individual SDRR series, and calculated the correlation between the obtained coefficients (the HRV decay rate *a* and the HRV intercept *b*) and CRF indices (namely maximum VO2, maximum aerobic power, power at ventilator thresholds) using Spearman rank correlation. The robustness of this analysis has been tested by an extended sensitivity analysis (see “sensitivity analysis” subsection).

### Participants

The database used in this work consists of records of 18 young athletes (10 males and 8 females; 15.2 ± 2 year-old, see table 1)) of the Regional Physical and Sports Education Centre (CREPS) of French West Indies (Guadeloupe, France), belonging to a national division of fencing, or a regional division of sprint kayak and triathlon. All athletes completed a medical screening questionnaire, and a written informed consent from the participants and the legal guardians was obtained prior to the study. The study was approved by the CREPS Committee of Guadeloupe (Ministry of Youth and Sports) and the CREPS Ethics Committee and performed according to the Declaration of Helsinki. A short summary of the physiological characteristics of the studied group is presented in Table 1.

**Table 1.** Physiological characteristic of the 18 participants. Values indicated are medians [Inter quartile range]. VO_2_max: maximum oxygen consumption; MAP: Maximum aerobic power; Peak HR: peak heart rate reached during maximal effort test; PVT_1_: power at the first ventilatory threshold; PVT_2_ power at the second ventilatory threshold.

| Variable (unit) | Value |
| --- | --- |
| Number | 18 |
| Age (years) | 15.0 [14.0, 16.8] |
| Sex (Male) | 12 (66.7 %) |
| Weight (kg) | 62.9 [54.3, 76.5] |
| Height (cm) | 174.0 [165.0, 182.0] |
| VO<sub>2</sub>max (mL/kg/min) | 36.5 [32.6, 41.8] |
| MAP (W) | 222.5 [177.5, 297.5] |
| Peak HR (beat/min) | 187.2 [183.3, 190.1] |
| PVT1 (W) | 105.5 [79.2, 143.0] |
| PVT2 (W) | 169.0 [141.2, 246.8] |
| sport | Fencing: 10; Kayak: 6; Triathlon: 2 |

### Graded effort test (GET) measurement

GET were performed at the end of the off-competition season. The subjects performed under the supervision of a doctor in sport medicine a GET on an SRM Indoor Trainer electronic cycloergometer (Schoberer Rad Meßtechnik, Jülich, Germany) associated to a Metalyzer 3B gas analyzer system (CORTEX Biophysik GmbH, Leipzig, Germany). The room was climatized and did not have external light to provide similar temperature, humidity, and light for each GET. The participants were instructed not to take alcohol, caffeine, nor to practice intense sport activities during the 24 hours preceding their GET. The GET consisted of a 3 min cycling period at 50 watts, followed by an incremental power testing of 15 Watts each minute until exhaustion. At the end of the test, measurements were prolonged during a 3 min period to record the physiological recovery of the athletes. They were during this period sitting on the cycle ergometer.

Cardiorespiratory parameters were recorded breath-to-breath all along the test session. The ventilatory thresholds 1 (VT1) and 2 (VT2) were calculated using the Wasserman method (22). Maximum aerobic power is the maximum power achieved during the last completed step of the GET. Peak HR and VO_2_max are the maximum values of the HR and VO_2_ averaged other 5 breaths. The RR series were derived from Electro Cardiogramm (ECG) recordings (Cardio 110BT, Customed, Ottobrunn, Germany, with 12 derivations). The time resolution of the ECG recording was 1 ms. Heart rate (HR) is calculated as

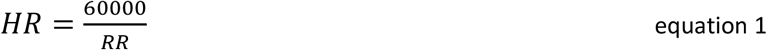

where RR is in millisecond and HR in beat/min.

### Data cleaning

Prior to performing the statistical analysis, we removed the artifacts in the RR series {*x*_1,_ *x*_2,_ …, *x*_*n*_} due to connection errors in the electrodes according to the following steps:

- All RR intervals with a value above 1000 ms during effort were removed. This concerned 0.02% of the RR measurements.
- At each point, if the RR value exceeded 2 times or was inferior of the half of the median value of the RR calculated in a 201 RR values windows centered on *x*_*i*_, it was removed. This concerned 0.02% of the measurements.
- At each point *x*_*i*_, if the absolute RR change *x*_*i*_ − *x*_*i*−1_ exceeded 10 times the median value of the RR increases calculated in a 201 RR values windows centered on *x*_*i*_, the point was removed. This concerned 0.5% of the measurements.

An example of points cleaned in an original raw RR series is presented in Fig 1.

**Fig 1.**
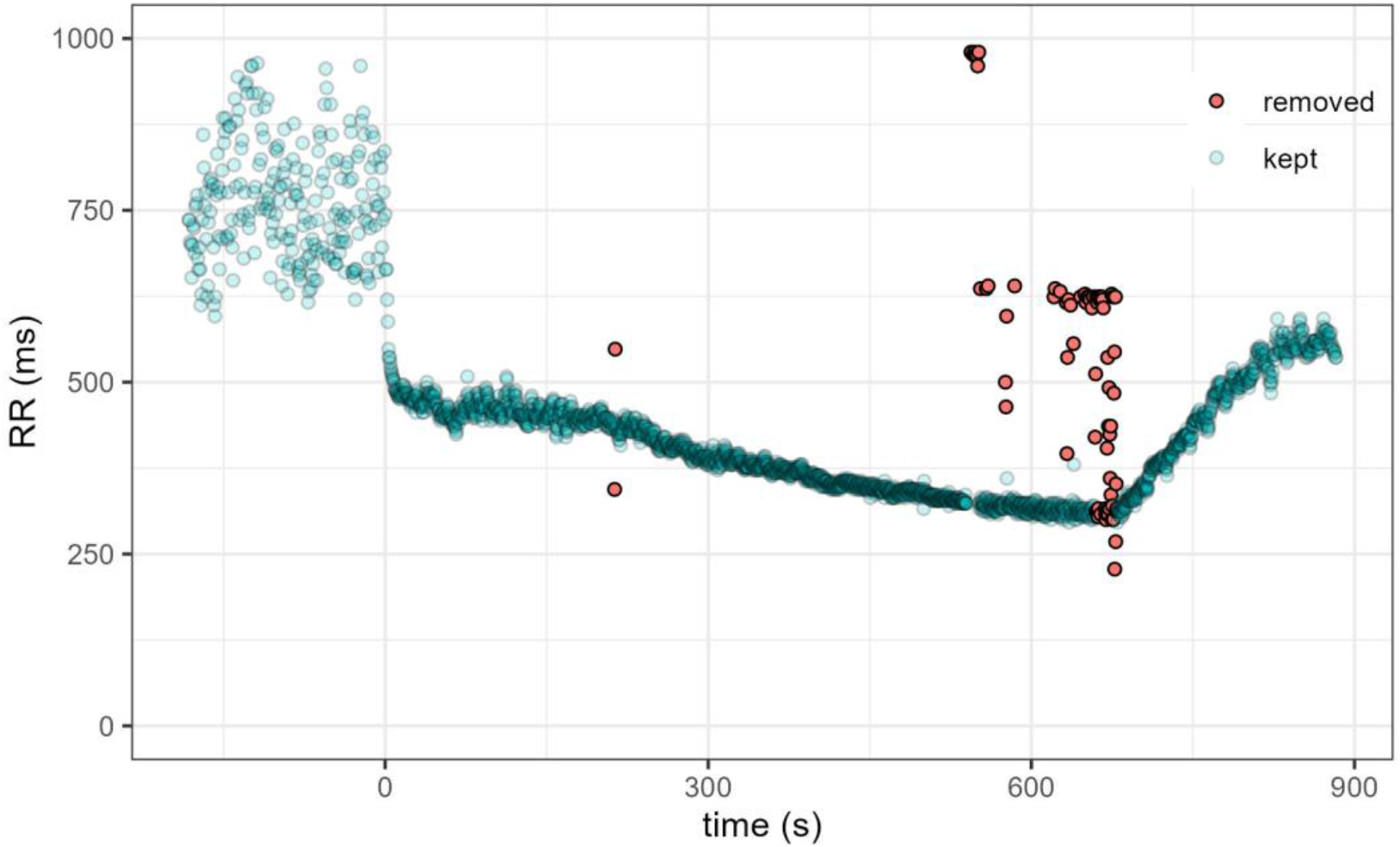
Measured RR interval during a maximal graded effort test (effort starts at time = 0). The points removed by our cleaning procedure are indicated in red

### Detrending

We used recent developments in dynamical analysis to model the non-stationary aspect of HR. The main trend of HR dynamics during a GET can be modeled by a simple first order differential equation driven by the power expenditure. A two-step estimation procedure, consisting in first estimating the time derivative of HR using a spline regression and then obtaining the constant coefficient of the differential equation through a linear regression, produces unbiased estimation of the differential equation parameters (23). An estimated curve can then be produced by numerical integration of the differential equation with the obtained coefficients. This simple model produces indices sensitive to CRF and performance changes (24). The possibility to estimate the gain (the amplitude of the HR increase corresponding to a workload increase) for each power step of the exercise test allows to reproduces up to 99% of the HR dynamics during the GET, and yields coefficients varying consistently with the metabolic changes associated to the respiratory thresholds (25). Because of the absence of ad-hoc parameters and the fact that it has been theoretically and practically validated, this procedure to estimate HR during exercise was used in the present study to detrend the RR time series.

### Heart rate variability

Given a series of *n* detrended RR intervals {*x*_1,_ *x*_2,_ …, *x*_*n*_}, the series of standard deviations of the RR intervals SDRR calculated on a successive window of *ω*_*RR*_ RR intervals is

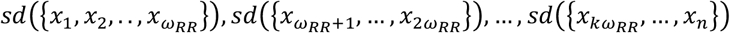

Where *sd* is the standard deviation, and *k* the integer part of *n*/*ω*_*RR*_. This calculation has the advantage of its simplicity, thus being easily reproducible. We have considered a window size of 1 min for the SDRR calculation in the main study in order them to correspond to each power step during the exercise test.

Because HRV is a result of parasympathetic and sympathetic neuronal activity, both being processes taking part in the regulation of the mean HR, we propose to analyze the decay of SDRR during exercise as a function of the mean HR calculated on the same time windows.

### Sensitivity analysis

In order to test the sensitivity of our results to the approach proposed, we performed a sensitivity analysis by:

– testing more classical polynomial detrending methods of different orders (26) to obtain stationary RR time series, instead of our parameter-free approach based on HR dynamical model;
– varying the windows size *ω*_RR_ used for SDRR calculation.

The polynomial detrending procedure can be described as follows: let us consider a time series of RR intervals and let be *ω*_*p*_ (odd integer) the window size and *p* the polynomial order. For each value of the RR series {*x*_*i*_}, we perform a least squares polynomial regression of order *p* on the data inside the window of size *ω*_*p*_ centered at position *i*. The detrended value at position *i* (*x*_*det,i*_) is then obtained by subtracting the estimated value produced by the polynomial regression 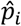 to the experimental value *x*_*i*_:

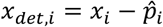

During the sensitivity study, the following parameters have been varied:

– The degree *p* of the polynomial has been set to *p* = 0, 1 and 2 (respectively equivalent to substract the mean, the linear fit and the quadratic fit inside the window).
– The window size of the local polynomial regression *ω*_*p*_ has been set to odd integers between 5 and 101.
– The window size *ω*_*RR*_ used to compute the RR time standard deviation SDRR has been varied considering the same range, i.e. odd integers between 5 and 101.

We thus tested the robustness of our analysis for 49×49×3 = 7203 different evaluations of SDRR change during effort for each of the 18 subject’s RR measurements.

### Statistical analysis

All signal processing and statistical analysis have been performed with the R 4.1 open source software (27). The comparison between regression models is based on Akaike information criterion (AIC) and Bayesian Information criterion (BIC). The calculation of the correlations between the estimated HRV decay coefficients and the CRF indices has been performed using Spearman rank correlation coefficients *ρ*. The regression of the three models proposed in equation is operationalized with a nonlinear least square regression using the packages *nls*. The *ggplot2* was used for graphical representation (28) and *data*.*table* for data manipulation.

## Results

The median number of RR distance recorded was 2726 beats (Inter Quartile Range [2433;3548]). An example of detrended RR estimated using the dynamical model presented in the method section is illustrated in Fig 2. This model produced an individual estimation of RR with a median R2 of 0.97 [IQR: 0.91 – 0.98].

**Fig 2.**
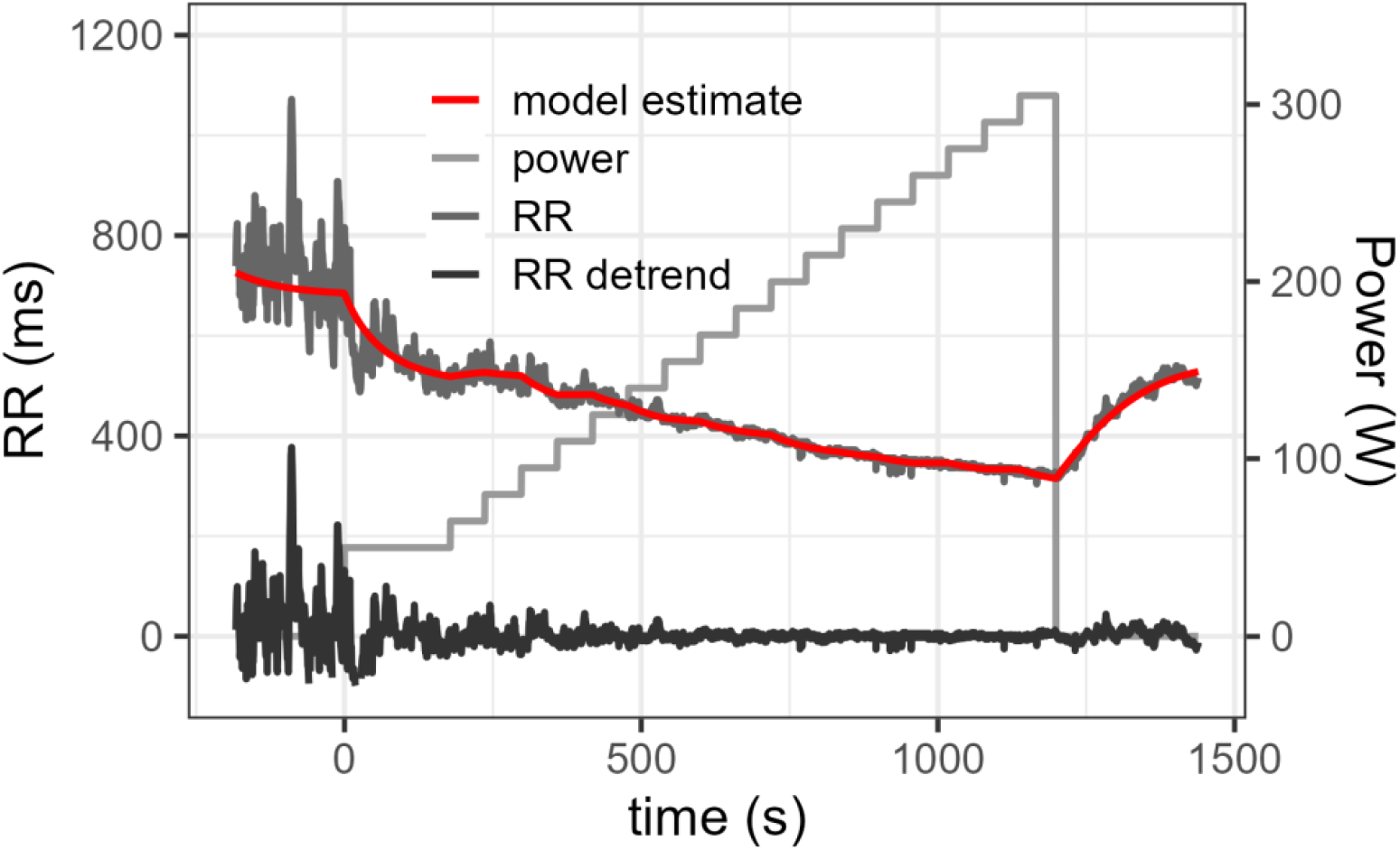
Measured RR interval during a maximal graded effort test, its estimation by a first order differential equation and the resulting detrended RR values obtained by subtracting the model estimate (2) to the real data.

### Comparison of models

In this first part we want to determine which representation of the evolution of SDRR along exercise correspond the best to an exponential decay. The ensemble of the SDRR values along the GET of all 18 participants are represented in Fig 3 on a logarithmic scale as a function of the corresponding mean HR (Fig 3A), as a function of the corresponding mechanical power exerted during the exercise test (Fig 3B) or as a function of the corresponding work intensity (Fig 3C). When plotted as a function of HR (model 1, Fig 3A), the ensemble of SDRR measured before, during and after the exercise align nicely on a linear-log scale, indicating a clear exponential behavior. When displayed as a function of the mechanical work rate (Fig 3B, model 2) as proposed by Lewis and co-authors (11), the values of HRV at high work rate do not align with the rest of them in a linear-log scale. Furthermore, the HRV calculated before and after the effort have a wide variety of values for the same null power. Finally, when represented as a function of %VO_2_max, although SDRR values plotted in linear-log scale display a clear linear trend during exercise, they do not align with the values calculated before and after effort (Fig 3C model 3).

**Fig 3.**
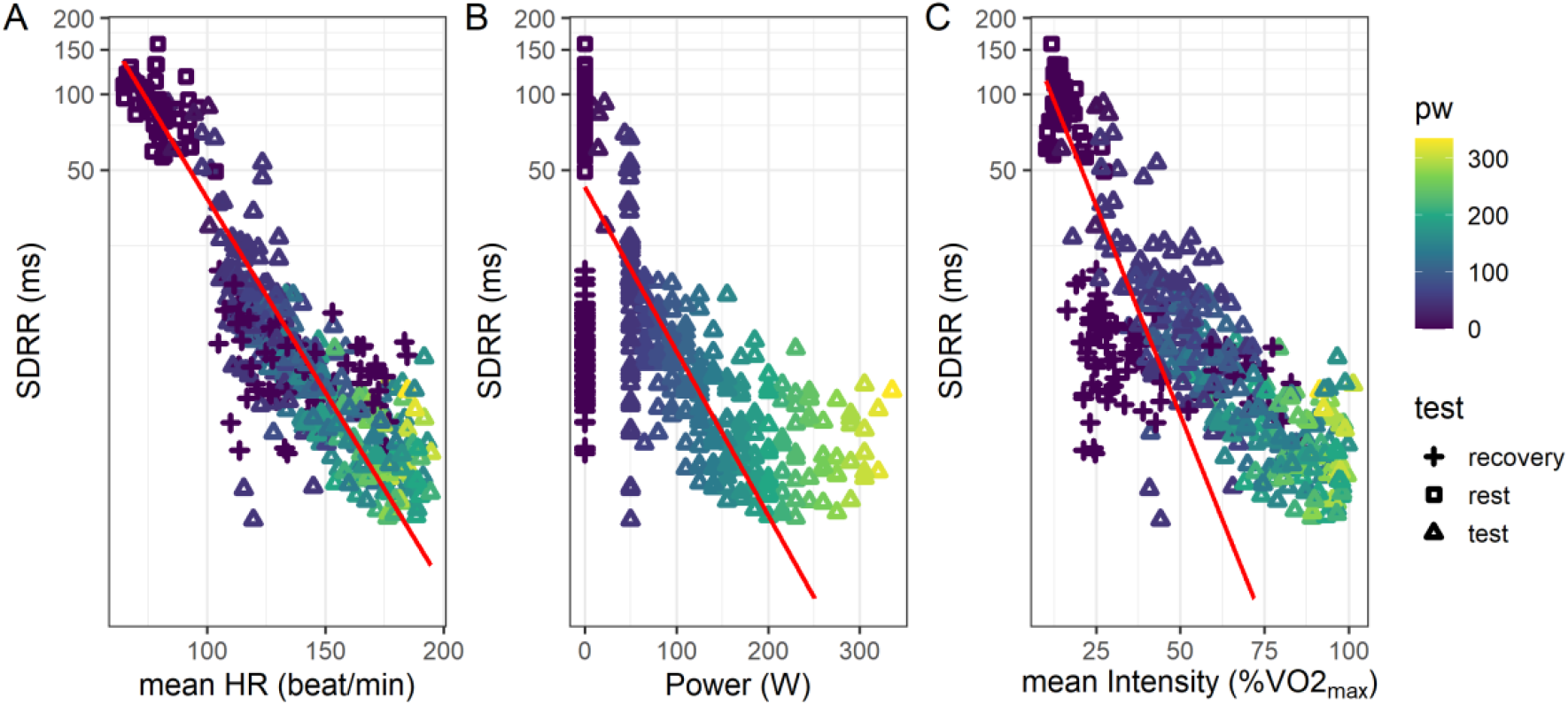
SDRR of the detrended RR time series recorded during a graded effort test as a function of: A) the mean heart rate (HR) during the interval, B) the mechanical power during the exercise test and C) the mean work intensity. The shape of the points indicates the SDRR was calculating on RR during (effort), before (pre) or after (post) the effort. The red line corresponds to the regression estimate obtained using model 1, 2 and 3 of equation 3 in A, B and C respectively

When performing the least square nonlinear regressions of these three models on the entire dataset (the estimated model is plotted as a red line in Fig 3), the model 1 in Equation 2 describing SDRR as an exponential decay of the mean HR has a significantly lower AIC and BIC than the two others, both when considering cardiac measurement only during exercise or when including HRV before, during and after exercise (see table 2).

**Table 2.** Akaike information criterion (AIC) and Bayesian Information criterion (BIC) for the models 1, 2 and 3 in equation 2 when applied to the ensemble of the SDRR computed on different part of the exercise test: before test (pre), during test (effort) or during recovery (post).

| model | effort range | AIC | BIC |
| --- | --- | --- | --- |
| model1 | pre + effort + post | 3179 | 3191 |
| model2 | pre + effort + post | 3855 | 3867 |
| model3 | pre + effort + post | 3477 | 3489 |
| model1 | effort | 1976 | 1987 |
| model2 | effort | 2080 | 2091 |
| model3 | effort | 2068 | 2079 |

The mean coefficients of model 1 in equation 2 estimated on the ensemble of the detrended SDRR series are *b* = 1325 ms and *a* = 0.035 min.beat^-1^ (*p <* 0.0001 for both coefficients), meaning that for young athletes, an increase of 10 beat/min of HR divided SDRR by exp(0.5)= 1.41, i.e. decreased it by ∼30% on average.

### Study of individual decrease of HRV during effort

Each individual SDRR series had a median of 22 measures [IQR 20-28]. The correlations between the individual parameters *a* (the HRV decay rate) and *b* (the HRV intercept), obtained when performing the nonlinear regression of model 1 on each detrended RR series, and the aerobic performances indexes, are reported in table 3.

**Table 3.** Spearman correlation coefficients *ρ* with the associated p value between the estimated HRV decay coefficients (HRV decay rate *a* and the HRV at HR=0 *b*) and aerobic performance indices: maximum oxygen consumption (VO_2_max), maximum aerobic power (MAP), maximum experimental heart rate Peak HR power at the first and second ventilatory threshold (PVT_1_ and PVT_2_) and Heart rate recovery (HRR)

|  | **a** |  | **b** |  |
| --- | --- | --- | --- | --- |
| | Spearman correlation $\rho$ | P value | Spearman correlation $\rho$ | P value |
| **VO<sub>2</sub>max** | 0.59 | 0.009 | 0.60 | 0.009 |
| **MAP** | 0.60 | 0.009 | 0.64 | 0.004 |
| **PVT<sub>1</sub>** | 0.53 | 0.02 | 0.58 | 0.01 |
| **PVT<sub>2</sub>** | 0.50 | 0.03 | 0.54 | 0.02 |
| **Peak HR** | 0.04 | 0.88 | 0.11 | 0.66 |
| **HRR** | -0.46 | 0.05 | -0.54 | 0.02 |

The HRV decay rate ***a*** of SDRR as a function of HR is strongly and significantly correlated with VO2max (ρ = 0.6, p < 0.009), with the maximum power reached during effort test MAP (ρ = 0.6, p < 0.009), and with the power at the ventilatory thresholds (ρ = 0.50, p = 0.03 and 0.5, p = 0.02 for PVT1 and PVT2). The HRV intercept *b* is also strongly positively correlated with VO2max (ρ = 0.6, p = 0.009), with the maximum power reached during effort test MAP (ρ = 0.6, p < 0.004), and with the power at the ventilatory thresholds (ρ = 0.64, p = 0.005 and 0.64, p = 0.004 respectively for PVT1 and PVT2). This indicates that participants with better aerobic performance have a higher SDRR at low HR, which decays more rapidly as HR increases.

### Sensitivity analysis

In Fig 4, the correlation between HRV decay rate *a* and VO_2_max and the associated p value is represented for all tested *ω*_*p*_ and *ω*_*RR*_ values when using 0^th^ order local polynomial detrending. Within the red square (*ω*_*RR*_ > 50 and *ω*_*p*_ > 50), ∼90% of the analysis yielded significant correlation with value higher than 0.5. The sensitivity analysis demonstrated that the previous results can be obtained using a simpler 0^th^ order local detrending with *ω*_*p*_ > 50 (i.e. performing the local polynomial detrending in windows of at least 50 points) to obtain a stationary RR series and calculating SDRR on the detrended RR series in windows of at least 50 points (*ω*_*RR*_ > 50). The results of the sensitivity analysis for other polynomial order and other CRF indices (ventilatory thresholds and maximum power) are given in supplementary material.

**Fig 4.**
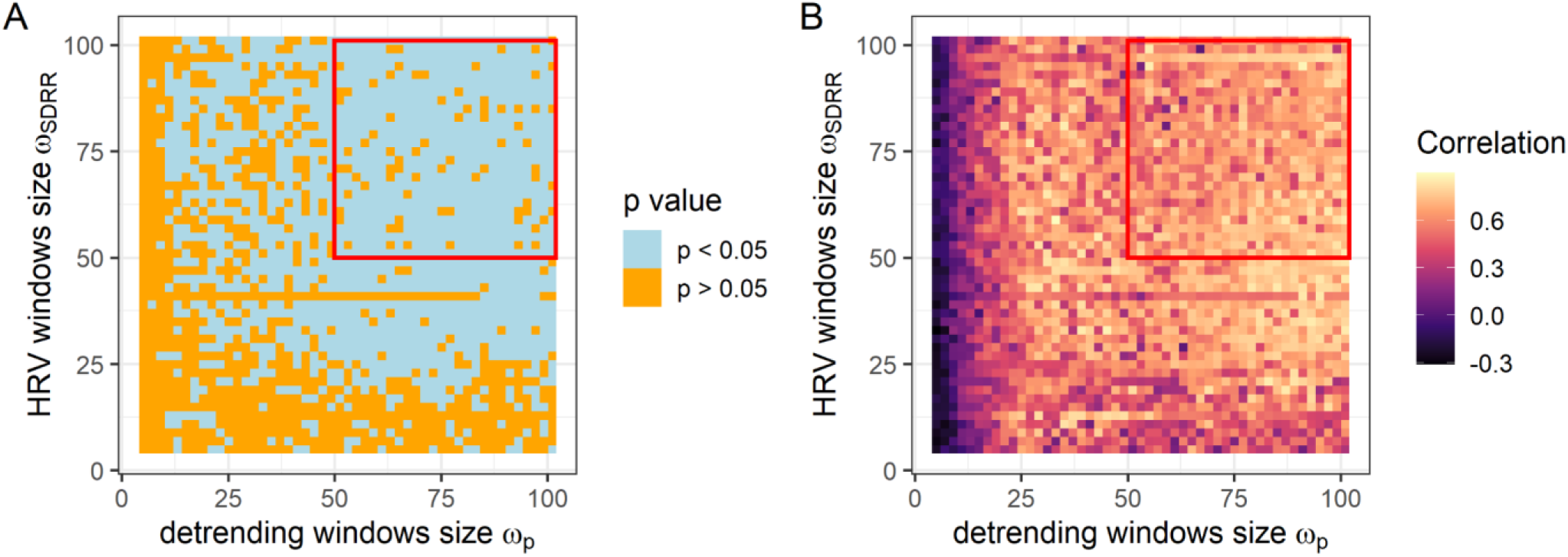
p value higher or lower than 0.05 (panel A) and Spearman correlation coefficient (panel B) between the HRV decay rate *a* and VO_2_max using a 0^th^ (p=0) order polynomial detrending. Each pixel corresponding to 1 of the 49 size of detrending window *ω*_*p*_= 1,3,5…101 and 49 sizes of SDRR calculation window *ω*_RR_=1,3,5. 101. Red squares delimit the regions for which we observe significance (p < 0.05) (A) and a high correlation (B) for 90% of the tests.

## Discussion

Our study aimed at studying how HRV varies during effort and how this evolution is linked with CRF. SDRR varies as an exponential decay of HR, and decreases by 30% every increase of 10 beats/min of HR. This fast decay causes time HRV indices to reach values close to the minimum allowed by the resolution of the ECG device early during the GET, thus explaining the absence of significant differences between HRV measured during constant load exercise at high intensities (14). On the other hand, the exponential decay rate of SDRR extracted by a nonlinear regression during an incremental exercise is strongly related to several parameters linked with aerobic performance, such as VO_2_max, MAP, and power at the ventilatory thresholds. This HRV decay rate increases (i.e. a faster decay of HRV when HR increases) for athletes with higher aerobic capacity. The analytical approach proposed only requires cardiac measurements and make use of measurements before, during and after the exercise.

This study is to our knowledge the first one proposing to observe the change of HRV during exercise as a function of the corresponding heart rate. Compared to the more common approach consisting in studying the change of HRV as a function of exercise intensity, our representation is better described by the exponential decay during exercise and unifies under the same model the HRV calculated on cardiac measurement before and after the effort. The similarity of these two representations during the effort resides in the linear relation that HR and VO_2_ have when performing graded exercise tests (29). Their differences reside in the shorter dynamical time of VO_2_ compared to HR, and is revealed at exercise onset and exercise cessation (24,30).

Previous studies reported higher HRV for trained subjects than untrained subjects at rest or at low exercise intensity (31,32). This result finds its roots in the higher vagal (parasympathetic) neuronal activity for trained subjects compared to untrained ones, as proved by the increase of vagal-related indices of resting and post-exercise HRV (33). Post-exercise HR recovery studies have also shown that trained subjects have a faster re-activation of their vagal activity at exercise cessation (34). The rate at which HRV decreases when the HR increases during activity is another measure of vagal activity, which has been shown to be directly linked with the exercise capacity (35), explaining thus the strong link between rate of HRV change during exercise and CRF. In our study, individuals with higher CRF start their exercise with a higher HRV due to their high vagal tone and have a faster subsequent decrease of HRV when their HR increases during exercise due to the faster withdrawal of their parasympathetic activity.

The strong correlation between our HRV decay characterization and performance indices found among a heterogeneous population of athletes in term of sport modality is a strength. Indeed, although these different sport modalities require unequal sources of energy and train distinct physical capacities, leading to different cardiorespiratory and cardio autonomic control features, the exponential decay of HRV as a function of HR seems to be a robust indicator of the cardiovascular fitness. On the other hand, the limited number of athletes in each sport category did not allow us to compare the changes of HRV during exercise between sport modalities.

The use of a physiologically motivated model of HR dynamics during exercise using no ad-hoc parameters to obtain the detrended RR series and the extensive sensitivity study associated shows the robustness of our approach and facilitates future similar studies by providing guidelines of necessary data acquisition and analysis methods used.

Nevertheless, the limited age range and physiological characteristics of the participants are limitations, and further studies are needed to generalize our results to a more diverse population. The cross-sectional aspect of our approach should be also complemented by a longitudinal approach.

## Conclusion

The present work demonstrates that the measure of the SDRR decay during exercise offers a solid index of physical capacities. Our study proposes a simple model to describe the changes of HRV with effort: SDRR behaves as an exponential of the heart rate. The characteristics of this exponential decay of HRV are highly dependent on the physical capacities and on the cardiorespiratory fitness. The proposed analysis, relying only on cardiac measurements and based on a set of simple mathematical tools, pave the way to the measurement of cardiorespiratory fitness using measurements provided by mobile devices.

## Data Availability

The data used for this study are openly available at https://doi.org/10.13026/7ezk-j442 All the code used to clean the data, perform the analysis, and to create the figures and tables is available in the Gitlab repository https://gitlab.com/dmongin/scientific_articles/-/tree/main/decrease_HRV_effort.

## Availability of data and material

The data used for this study are openly available at https://doi.org/10.13026/7ezk-j442.

## Code availability

The detrending method based on first order differential equation has been made freely available in the R package doremi: https://cran.r-project.org/web/packages/doremi/index.html. All the code used to clean the data, perform the analysis, and to create the figures and tables is available in the Gitlab repository https://gitlab.com/dmongin/scientific_articles/-/tree/main/decrease_HRV_effort.

## Authors’ contributions

Material preparation and data collection were performed by C.C. and O.H.. Methodology was designed by D.M., P.C. and P.A.B.G.. Analysis was performed by D.M.. Writing of the article, was done by D.M., M.G.E. and P.A.B.G., preparation of the Figs and tables was done by D.M.. All authors reviewed the manuscript and approved the present version of the work.

