## Supplementary material for "Decrease of heart rate variability during exercise: an index of cardiorespiratory fitness"

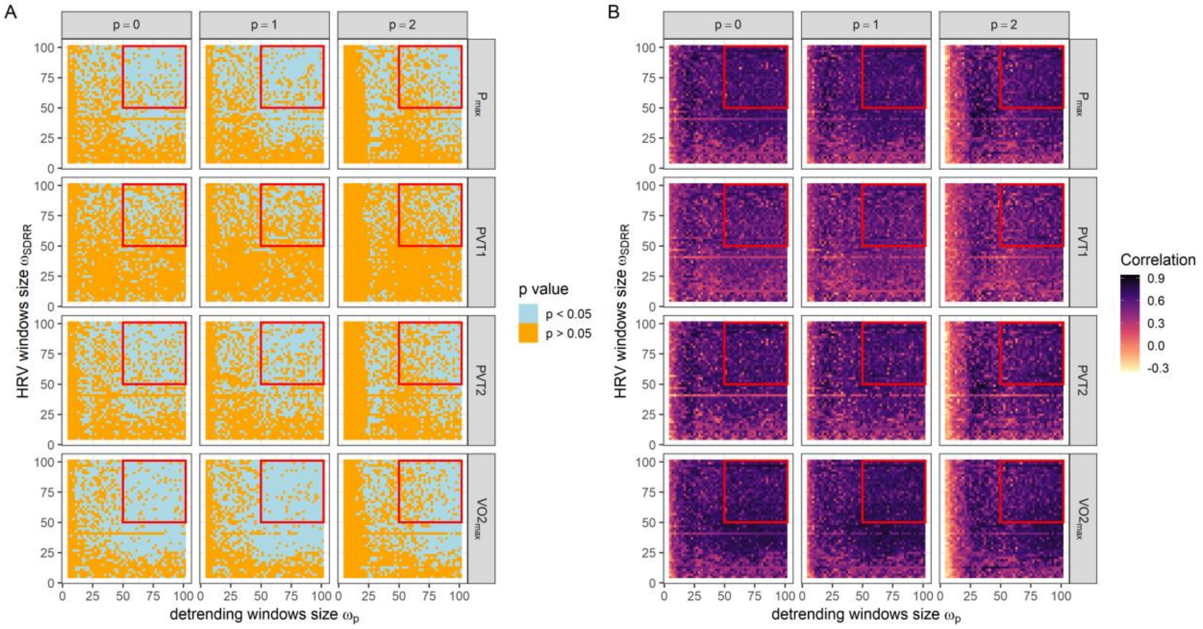


Figure S1. p value (panel A) and Spearman correlation coefficient (panel B) between the HRV decay rate a and cardiorespiratory parameters (Pmax, PVT1, PVT2, VO2max), using a 0th (p=0), first (p=1) or second (p=2) order polynomial detrending. Each pixel corresponding to 1 of the 49 size of detrending window $\omega p$= 1,3,5...101 and 49 sizes of SDRR calculation window $\omega RR$=1,3,5...101. Red squares delimit the regions for which we observe significance below 0.05 (A) and a high correlation (B).
